# Effectiveness of the strategies implemented in Sri Lanka for controlling the COVID–19 outbreak

**DOI:** 10.1101/2020.04.27.20082479

**Authors:** K.K.W.H. Erandi, A.C. Mahasinghe, S.S.N. Perera, S. Jayasinghe

## Abstract

In order to bring the new coronavirus pandemic in the country under control, the government of Sri Lanka implemented a set of control strategies including social distancing, quarantine, lockdowns, travel restrictions and isolation of villages. The aim of this study is to investigate the effectiveness of the overall control process with the aid of classical compartment models and network models. Our results indicate that the prevailing control strategies are effective with at least 50% contact rate reduction or with at least 40% isolation of the contact history of infected population.

## 1 Introduction

An outbreak of a novel coronavirus (COVID–19) was reported from Wuhan province, China in December 2019 [8,20], causing numerous deaths and complications such as pneumonia and acute respiratory distress syndrome [9,16,21]. The infection rapidly spread to all parts of the globe and was declared a pandemic on 11^th^ March 2020 by the World Health Organization [14].

Sri Lanka reported the first case in a Chinese tourist on 27^th^ January 2020 and subsequently in a local person on 11^th^ March, 2020 [19]. Aimed at controlling the pandemic, the government enforced a strict strategy of case detection, identification of contacts, quarantine, travel restrictions and isolation of small villages as well, changing the strategy in a timely manner, that has so far been successful in confining the epidemic to 2,810 cases as of 29^th^ July 2020 and no community case was reported in the country since 30^th^ April 2020. Figure 1 shows the variation of total cases and new cases reported up to 29^th^ July 2020.

**Fig. 1:**
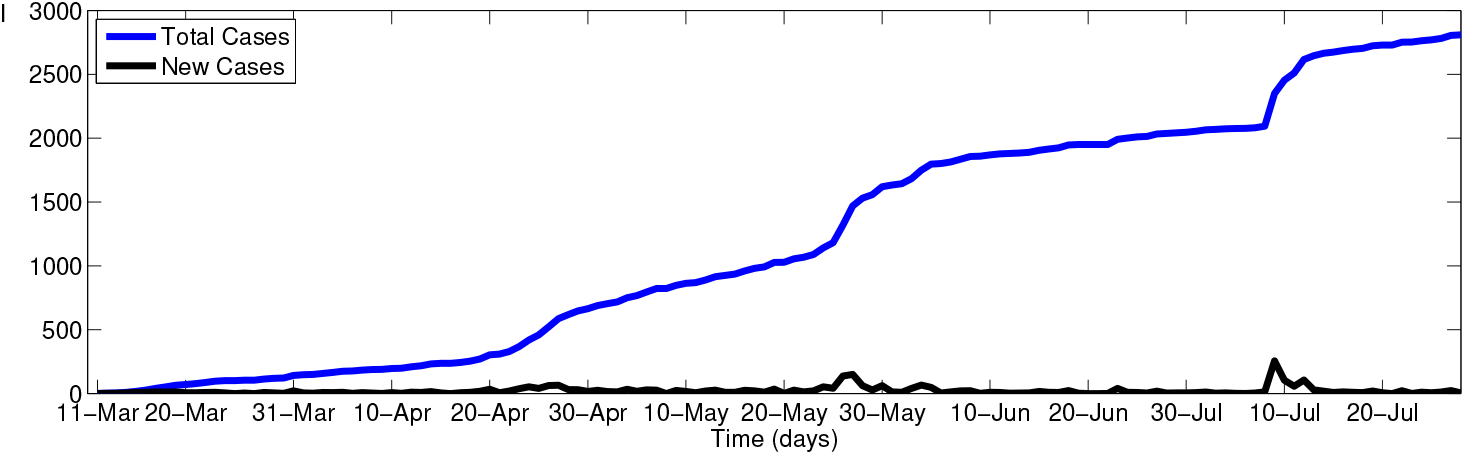
Reported COVID–19 cases from 11^th^ March 2020 to 29^th^ July 2020, Sri Lanka (Source: Epidemiology Unit, Ministry of Health, Sri Lanka)

Though some attention has been paid to other control measures such as spraying disinfectants, aimed at destroying the virus, the major control strategies implemented in Sri Lanka was aimed at minimizing the human movement. It is noteworthy that this was not a strategy used in the country even during the multiple epidemics of dengue, though others have long exercised restricting the mobility to control infectious diseases [2]. However, with the emergence of the COVID–19 outbreak, strict measures were taken to minimize human mobility. On 20^th^ March, a nationwide curfew was imposed till 24^th^ March and it was further extended together with a strict ban on inter–district travel. The curfew was a legal mechanism available to the newly elected government since parliament had been dissolved pending an election. Three populous districts of, Colombo, Kalutara and Gampaha were identified as high-risk zones. The main intention of the curfew was to implement a lockdown and minimize contacts with infected individuals, and thereby reduce the growth rate of the epidemic.

Entry of the virus to the island–nation was curtailed by closure of the airport and the ports together with the curfew. Extensive tracing was done of recent returnees from overseas (especially from China, Italy and South Korea) [17]. Identified patients were transferred to designated infectious disease hospitals and potential contacts to more than 40 quarantine centres, which were developed within a very short period and opened by the Ministry of Defense. In addition, on 21^st^ April 2020, the government began repatriating citizens stranded abroad and quarantine them for two weeks time period. In another example of quarantine, a whole village was sealed and quarantined in that location because an identified patient was found to have moved around 26 households in a circumscribed village. Entry and exit from the village was banned and they received free supplies and other amenities at the border. Starting from 26^th^ March more than 16 villages or defined small urban areas were sealed for 2–4 weeks in the country to control the spread of the disease. Moreover, on 22^st^ April 2020, a sailor from Welisara Navy camp was identified as a COVID–19 positive patient and immediately the camp was quarantined with 4,000 people at the Navy Camp. Since many sailors were on leave when contracted the disease, more than 6 villages were sealed off and around 1,300 people have been asked to self-quarantine.

Considering the progress of the control measures and the impact of the strict curfew to the economy of the country, the government has announced on 19^th^ April 2020 its decision to relax the nationwide curfew, and to implement moderated travel restrictions by permitting inter–district travels only to report for work and for essential services purposes. Further, on 11^th^ May 2020 the government had ordered partial opening of offices and businesses and the restrictions were further relaxed with a new set of rules. For an example, wearing a mask is mandatory in indoor and outdoor public spaces and public transport. The police and army personnel has been empowered to carefully scrutinize whether citizens are obeying with the rules. Moreover, the sanitary measures such as having wash basins and sanitizes outside the groceries and supermarket for customers has been made compulsory and the supermarkets were instructed to use thermometers to check the temperatures of the customers before they enter the premises. The distances are marked in the counter queues to minimize the close contacts. Schools however started academic activities only for grade 5, 11 and 13 on 6^th^ June and universities will continue online teaching and learning until further notice.

Almost all control strategies implemented in Sri Lanka were aimed at minimizing the human mobility, both the imposition and the relaxation of the curfew could be interpreted as changing the mobility rates in a certain way. From a policy perspective, it is relevant to know the extent of mobility that would be optimum to control the pandemic. This could be supported by reviewing the impact of mobility restrictions on the pattern of 254 reported cases up to 19^th^ April 2020 and comparing with the current situation. This would include the effects of curfew in some regions on other regions.

In order to review the impact of the control strategies implemented so far, we used the classical compartment models in epidemiology to model the transmission of COVID–19 in the three main districts. The restrictions imposed of human movement were incorporated by moderating the compartment model accordingly, by parametrizing contact rate and adding inter–district travel. Then we investigated the impact of relaxing the curfew by considering how the disease could be transmitted through the epidemiological network created by inter–regional transport.

## 2 Materials and Methods

In order to examine the transmission of the COVID–19, we adopted the *Susceptible–Exposed–Infectious–Recovered* (*SEIR*) model [1] with control measures applied to the high–risk zones in the country. Since the incubation period for the disease is 2–14 days as reported, there is a high possibility of transmission prior to the onset of symptoms and without showing symptoms [10], which justifies the adoption of the *SEIR* model. In the classical *SEIR* model, a population of size *N_i_* in the *i*th region is divided into susceptible (*S_i_*), exposed (*E_i_*), infected (*I_i_*) and recovered (*R_i_*) components. We extended the model by including an extra compartment as *hospitalized* (*H_i_*). This included all those entering hospitals and quarantine centers. The susceptible class consists of individuals who can possibly get infected by the disease. If the susceptible person is exposed to the virus, the individual is moved to the exposed compartment at the rate of *β_i_*. Let *α* be the transition rate of the exposed individuals to the infected class. Infected individuals, those who are hospitalized move to the newly added compartment *hospitalized* at the rate of *h*. Infected persons in compartments *I_i_* and *H_i_* move to the recovered compartment after the infectious period *γ*^−1^ and the persons in the recovered class are assumed to have permanent immunity against the disease.

For this study it is assumed that the mean infectious period is, *γ*^−1^ = 10.25 days [7]. Since COVID–19 virus load in the upper respiratory tract peaks within the first week of infection and transmission of the disease can be occurred 1–3 days before symptom onset, it is assume that the exposed person is infectious and able to transmit the virus to another person after 5 days of latent period (*α*^−1^) [13,12]. In order to scale *β_i_*, we used *fminsearch* in MATLAB.

Let *L_ij_* denote the percentage of daily travels from district *i* to *j*. We assumed that the percentage of daily inbound and outbound travel frequency to Colombo district is 15% and the percentage of daily travel frequency between Kalutara district and Gampaha district is 5%. Moreover, we introduced a parameter *contact rate*, symbolized by *c*, to quantify the degree of social distancing. We considered *c* as a normalized version of such a parameter; therefore the values of *c* lie between 0 and 1, and the case *c* = 1 is associated with a situation where no control measures are taken. A varying *c* represents the changing levels of control measures and it is assumed that the contact rate within each stage are relatively stable.

In order to investigate the impact of relaxing the curfew to other regions, we considered an epidemiological network in the form of a weighted graph, of which the vertices represent regions and the edges their inter–connections. Vertex weights were assigned proportional to the reported COVID–19 in the regions and the edges were weighted by considering the level of inter–connections, once the travel is permitted. It was assumed the inter–regional travel after relaxing the curfew resembles the travel before the lockdowns. Accordingly, highly vulnerable regions were identified, which needs to be paid more attention when future control measures are designed by Sri Lankan health–planners.

### 2.1 Transmission before the lockdowns

We first examined the transmission dynamics before the imposition of curfew. This situation is governed by the epidemic model with neither control measures nor quarantine. As stated previously, we consider the transmission in each district, the rate change of susceptible (*S_c_*), exposed (*E_c_*), infected (*I_c_*), recovered (*R_c_*) and hospitalized (*H_c_*) populations in Colombo district can be described by the following set of differential equations, where similar sets of differential equations can be built for Kalutara (*k*) and Gampaha (*g*).

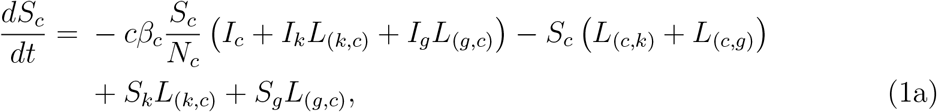

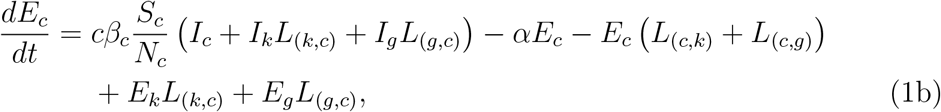

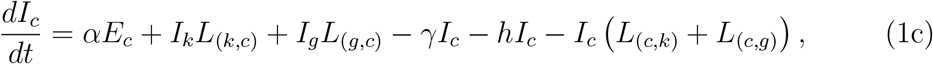

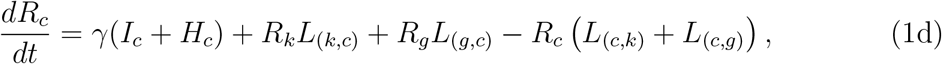

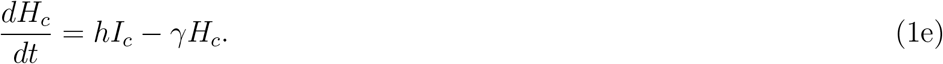

### 2.2 Transmission during the lockdown period

The situation during the lockdown which started from 20^th^ March was examined under three stages. The first stage is the period that starts from the first reported incidence to the date a curfew to a certain extent was imposed. Naturally, transmission dynamics has changed from this particular day, Thus, the contact rate reduced from this point on. To determine the contact rate reduction from 20^th^ March, we considered the difference between actual infected data and simulation data for several *c* values, as depicted in Figure 2 and based on the results c was set to be 0.8 from 20^th^ March. This period is considered the second stage. The third stage was when a strict inter–district travel ban was imposed from 24^th^ March onwards. Thus, the parameters needed to be changed again from this point on: that is, L*_i,j_* = 0 for all districts.

**Fig. 2:**
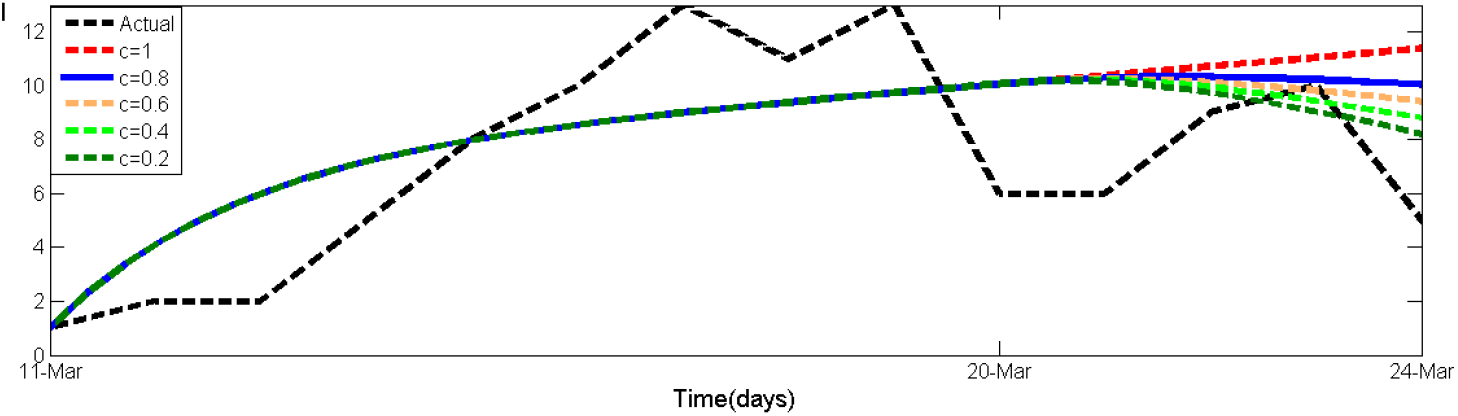
Comparison of actual infected data and simulation results for different contact rates in Sri Lanka from 20^th^ March to 24^th^ March

Also we considered the contact tracing took place during this period. During the lockdowns it was more convenient for the authorities to find the contact history of the infected individuals, and much progress seemed to be made in this regard. Based on the findings, the high–risk village was sealed in Kalutara district on 26^th^ March. Subsequently, several other small geographical regions were sealed–off. As a consequence, the number of self–quarantined individuals increased from 26^th^ March onwards. Hence, we added a new compartment named *quarantined* (*Q_i_*) for each district *i* in the existing model. The compartment *Q_i_* consists of exposed individuals identified by contact tracing and susceptible individuals living in the sealed–off villages. By enforcing contact tracing, *q* percentage of individuals exposed to the virus is quarantined. The quarantined individuals leave this compartment at a rate of *λ*, and move to the hospitalized compartment with a proportion *b*, if confirmed to be infected by COVID–19; otherwise, they go back to the susceptible class after the quarantined period of 14 days. The extended model that represents this scenario is expressible as follows:

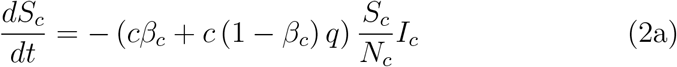

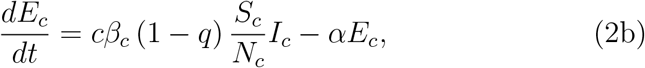

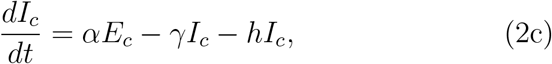

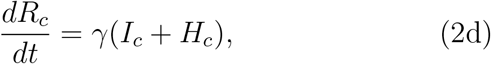

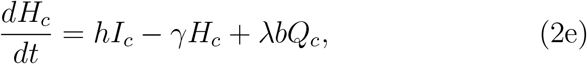

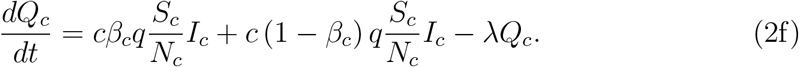

To analyze the effectiveness of physical distancing, we performed a sensitivity analysis on the parameter *c* and compared the simulation results with actual reported data.

### 2.3 Transmission after relaxing the lockdown

From 20^th^ April, the transmission dynamics of the disease changed again, as the government decided to relax the nationwide curfew. During this period the curfew was effective only between 8.00 pm to 5.00am and 50% of the workforce in state institutions were required to report to work. Travel between districts was permitted for purposes of employment or essential needs. Hence, we changed L*_ij_* values from 20^th^ April as the percentage of daily inbound and outbound travel frequency to Colombo district is 7.5% and the percentage of daily travel frequency between Kalutara district and Gampaha district is 2.5%. In addition, since all the citizens are ordered to follow the new set of rules and wearing face–masks other prevention strategies reduce the transmission risk by more than 50%, from 20^th^ April the transmission rate is reduced by 50% [4,5].

Moreover, to investigate the impact of curfew relaxation on each province in Sri Lanka, we considered the epidemiological network of provinces and the extended model for province *i ca*n be formulated as follows:

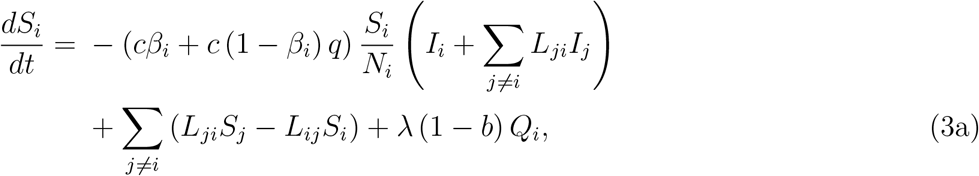

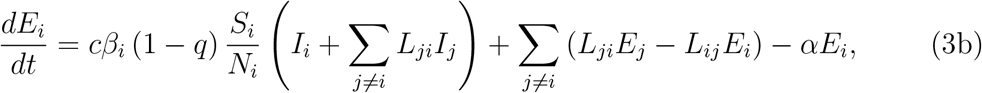

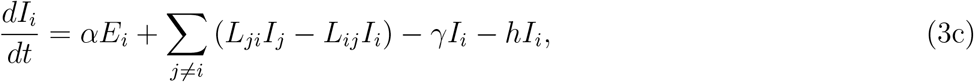

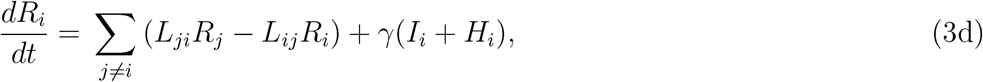

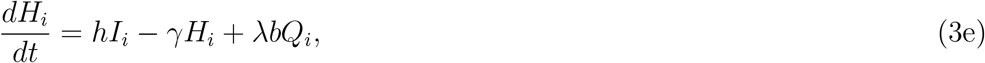

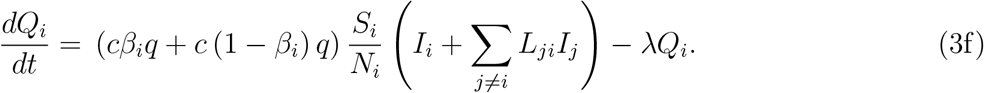

### 2.4 More consequences of relaxing

When the curfew was relaxed, public transport was allowed and the decision was questioned by several parties and was a subject of severe debate. Strict social distancing was followed in the country during the curfew, even in the waiting lines at chemists (which were opened to supply medicines during the curfew), supervised by the security forces. However, once public transport was made available, it was observed that the distancing was not followed. Therefore, the risk the virus being transmitted through the transportation network should not be ignored by the health planners.

Furthermore, it is important to examine the impact of public transport to the transmission of the virus. Assuming the public transport services operate at prevailing conditions, we determined which regions are most vulnerable, by considering a network model, of which the vertices represent regions and the edges their interconnections, weighted by numbers proportional to the number of public passenger vehicles available in those routes. Following [6], we too adopted the *closeness centrality*, defined by Equation (4), in order to measure the vulnerability of a province.

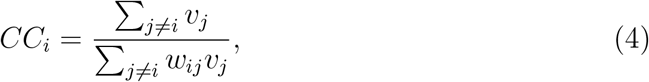

where, *υ_j_* denotes the total number of infected population as a faction of total population in *j*th province and *w_ij_* denotes the interconnection between province *i* and *j* in terms of human mobility.

We referred the COVID–19 situation reports available from the Epidemiology Unit, Department of Health, Sri Lanka [19]. We used the average number of closed contacts of reported infectious as the initial exposed number for the model simulation and assumed all of them were reported from Colombo district. That is, *I_C_* = 1 and *E_c_* = 26 at *t* = 0. In order to solve the developed model numerically, fourth order Runge–Kutta method was used with MATLAB. Table 1 represents the parameter values use in the model simulation.

**Table 1:**
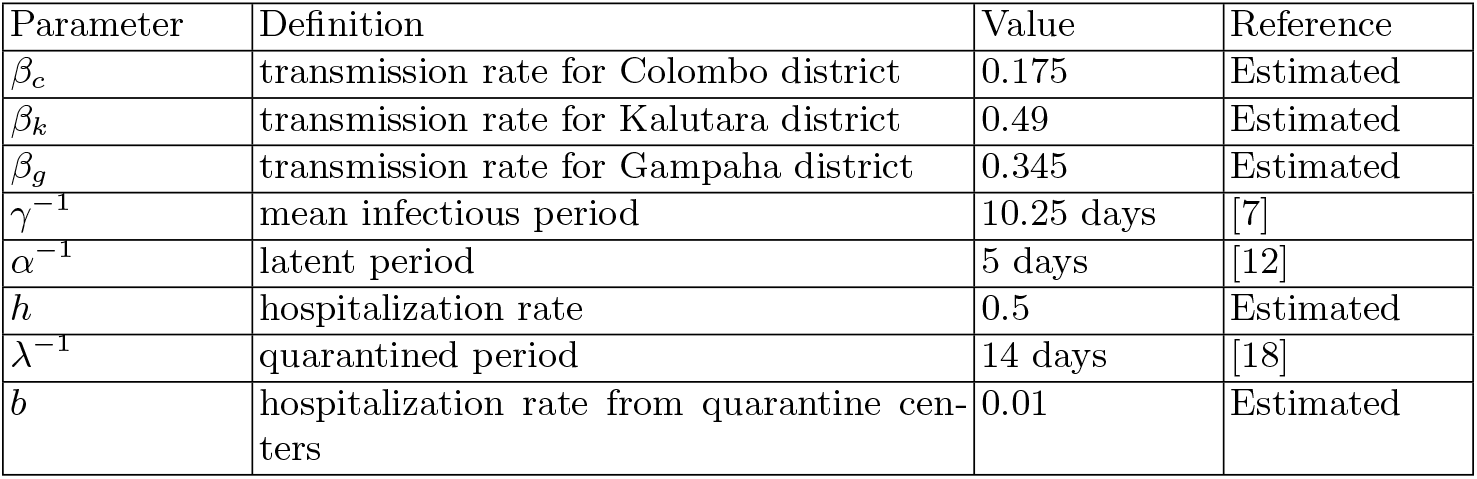
Descriptions and values of all parameters used in the model simulation

## 3 Results and Discussion

Figures 3, 4 and 5 illustrate the comparison of actual data and estimated infected population for different contact rates in Colombo, Kalutara and Gampaha districts respectively. There is a discrepancy between the actual data and simulation results in Figure 3, 4 and 5. As stated in the previous section we used the *fminsearch* in MATLAB and set up the *β_i_* for each district *i*. Therefore, each district obtained non–identical initial transmission rate values. Not surprisingly, the lowest *β_i_* value obtained for Colombo district; most populated district in the country and the highest *β_i_* value for Kalutara district; least populated district among the considered districts.

**Fig. 3:**
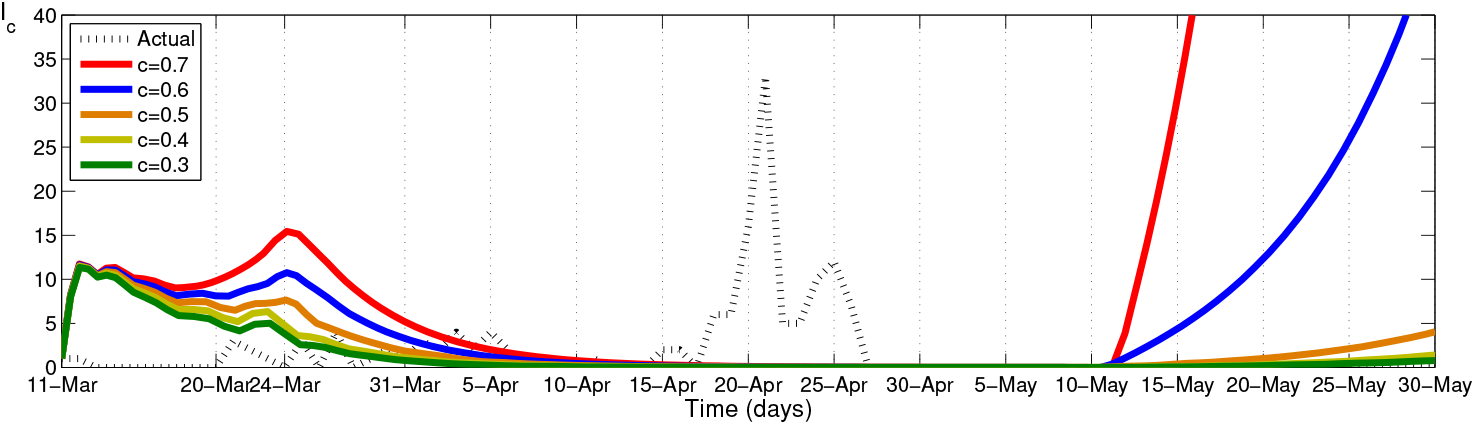
Comparison of actual infected data and simulation results for different contact rate values in Colombo district

**Fig. 4:**
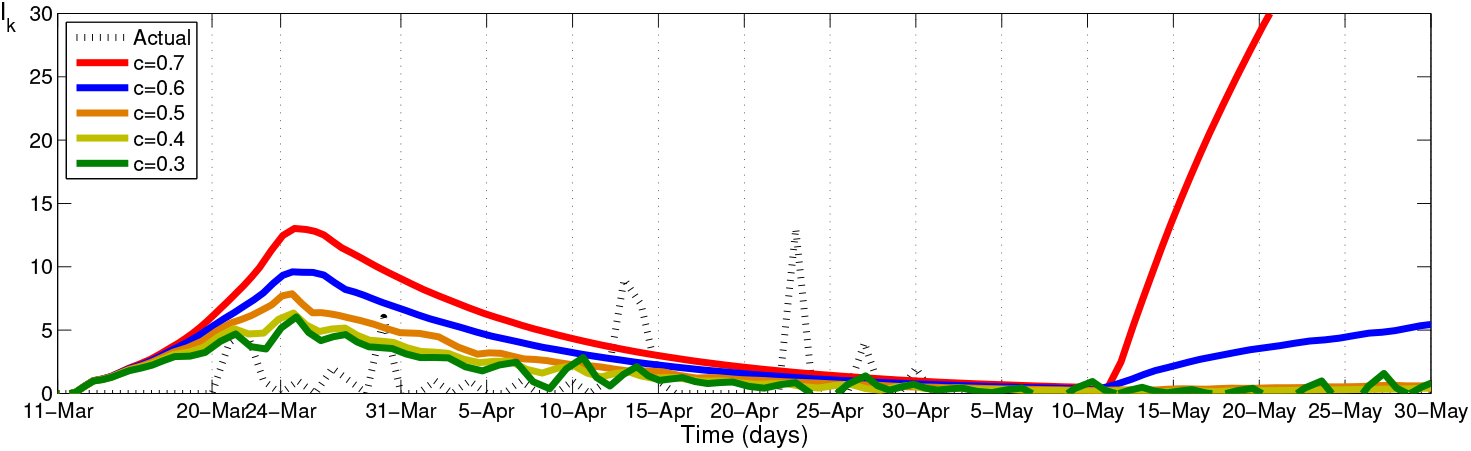
Comparison of actual infected data and simulation results for different contact rate values in Kalutara district

**Fig. 5:**
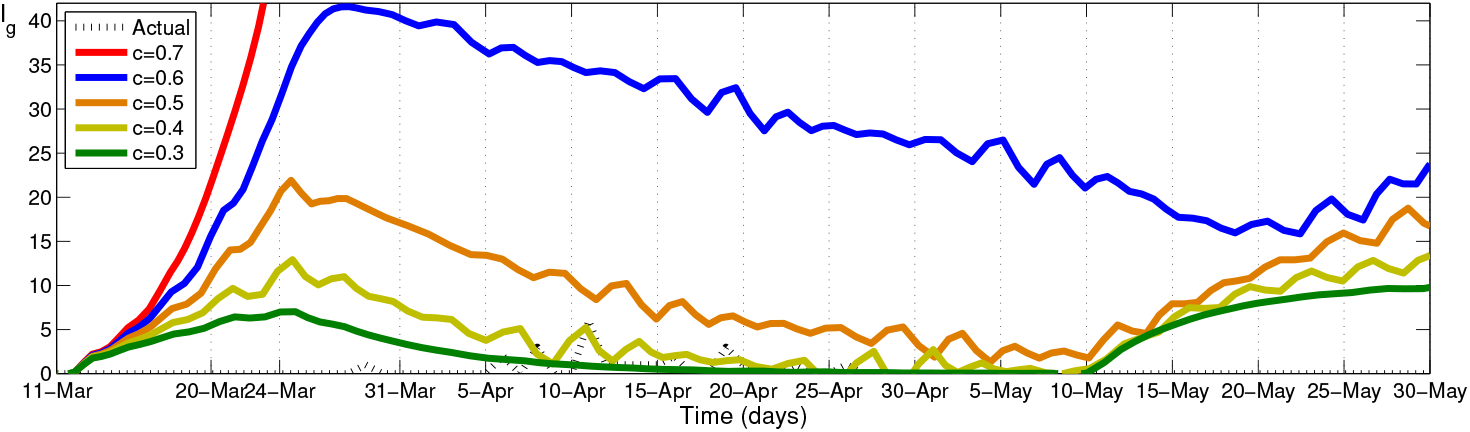
Comparison of actual infected data and simulation results for different contact rate values in Gampaha district

Therefore, the contact rate variation behaves differently for each province, hence the dynamical behaviour of infected population for different districts are significantly different as shown by Figure 3, 4 and 5.

It is clear from the simulation results in Figures 4 and 5, individuals in Kalutara and Gampaha districts get infected, even without any initially infected or exposed persons in the district. This indicates the significance of inter–district mobility in the transmission of disease.

To learn about the nationwide disease transmission, we simulated the model for the entire country by changing model parameters and compared the simulation results with the reported infected data. Figure 6 illustrates comparison of actual infected data and simulation results and Figure 7 represents the estimated exposed population in Sri Lanka for different contact rate values. From Figure 6, it can be observed that significant number of cases were after 30^th^ April and according to the Epidemiological Unit data all of them were reported from quarantine centers.

**Fig. 6:**
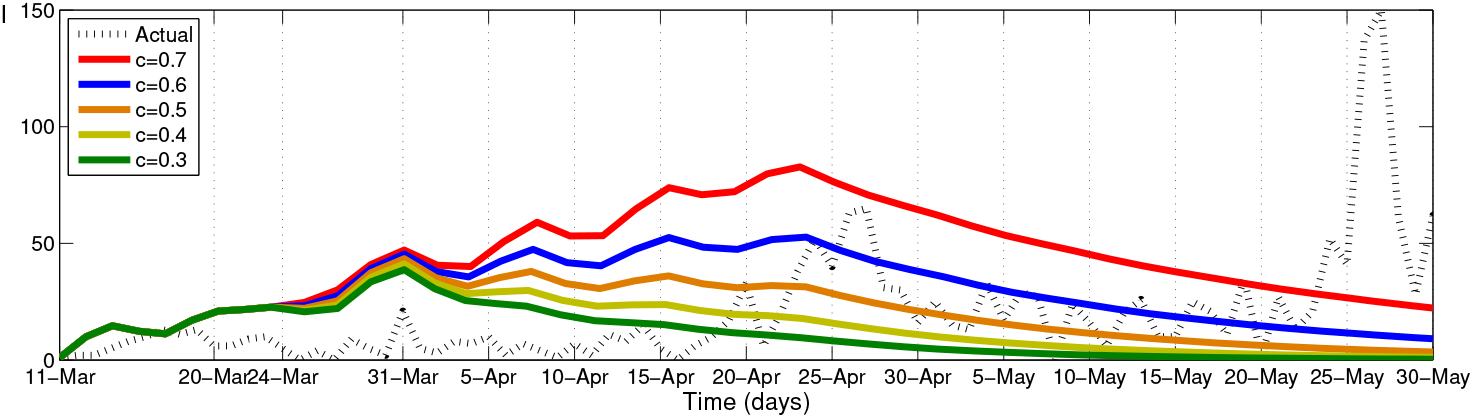
Comparison of actual infected data and simulation results for different contact rate values in Sri Lanka

**Fig. 7:**
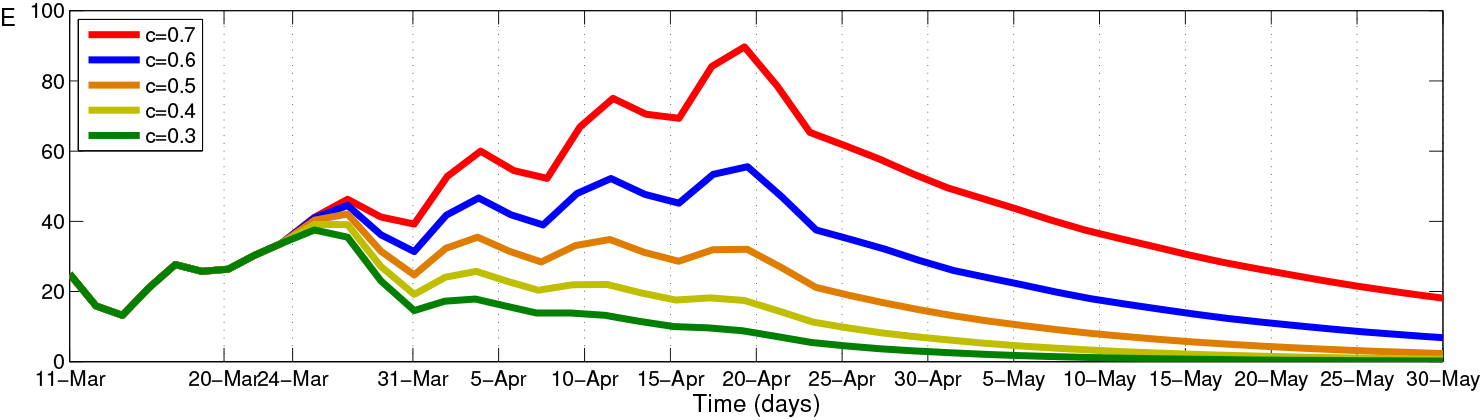
Estimated exposed population for different contact rate values in Sri Lanka

Computational results for several contact rate values suggest that the currently practising control strategies would be beneficial if the contact rate is reduced by not less than 50%. Moreover, it can be observed that the number of infected population could be reduced by increasing the number of control strategies.

In addition, to determine the effect of contact tracing and quarantine on the control of the spread of COVID–19, we performed a sensitivity analysis on parameter *q*. For the simulation, we assumed that contact rate is a constant value (*c* =0.8) from 24^th^ March. Figure 8 illustrates a comparison of actual infected data and simulation results for different quarantined percentage in entire country and the simulation results indicate the control of the disease spread could be done by tracing not less than 40% of the contact history of infected individual and quarantined them.

**Fig. 8:**
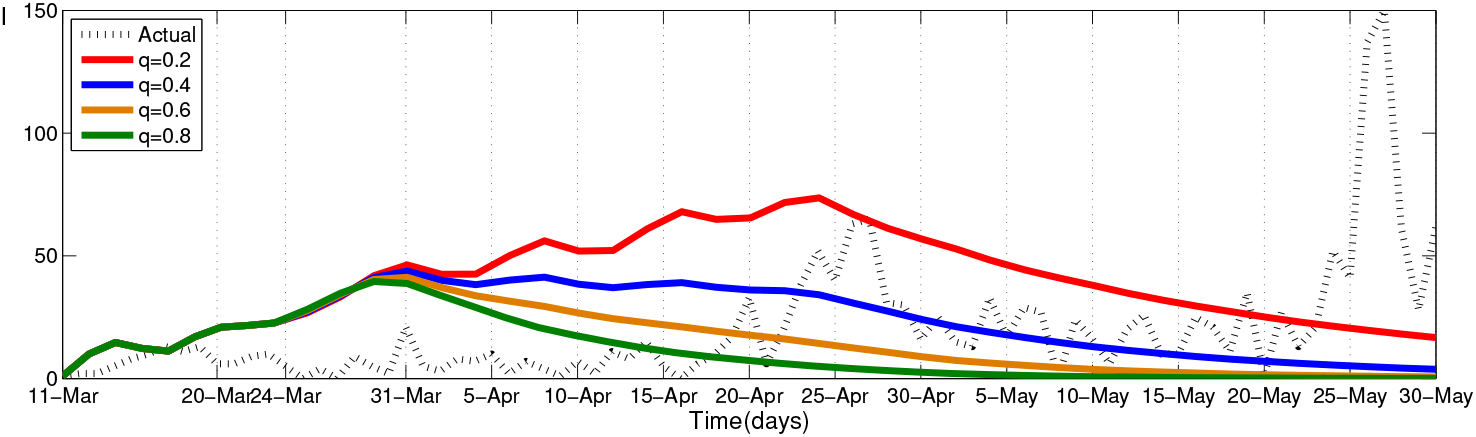
Comparison of actual infected data and simulation results for different quarantine values in Sri Lanka

Considering the less chances of getting admitted to a hospital and the possibility of having unidentified cases in the country cannot be ruled out in this context. According to the data of Epidemiological Unit, Sri Lanka ⅔ of infectious have no symptoms. In this sense, confirmed cases are only a fraction of the total infected population. Therefore, we believe the actual number of infectious persons is not exactly same as the number of reported cases on that day. According to recent studies [11,15] and WHO reports [14], 50% – 80% of infections are mild or asymptomatic. From Figure 9 it can be observed that the cumulative number of reported infectious cases range between 1/2 and 3/4 of estimated infectious cases upto 25^th^ April.

**Fig. 9:**
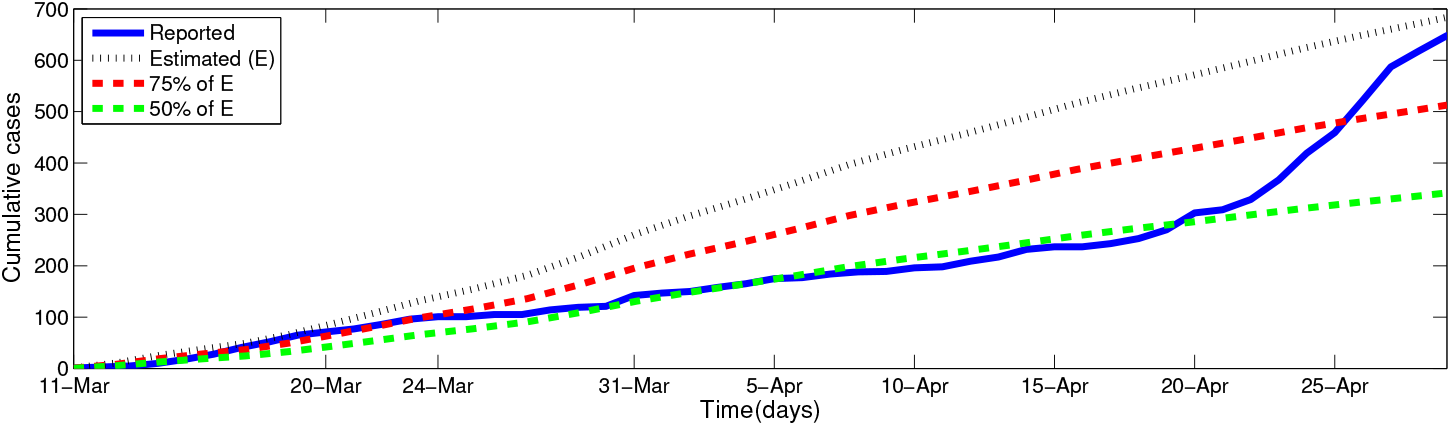
Comparison of actual infected data and simulation results for different percentage of *ρ* in Sri Lanka. (*ρ* is ratio of symptomatic to total infectious cases)

Recall that after 25^th^ April most of the identified cases were from navy Welisara Navy Camp cluster and most of them were asymptomatic cases. Therefore, after 25^th^ April difference between the number of reported cases and actual infectious cases was reduced. Since no community case was reported in the country after 30^th^ April 2020, we have done the comparison for the period from 11^th^ March to 30^th^ April 2020.

After identifying COVID-19 positive patient who is a sailor from Welisara Navy camp, the camp was quarantined with immediate effect with sailors and their families. When contracted the disease, many sailors were on leave and immediately all sailors were recalled to the camp and identified the contact history of each sailors. In a such situation, some villages or small regions have been sealed and residents have been asked to be self–quarantined based on the contact history of infected individuals. Once the high–risk regions are identified, the next immediate problem is how long these areas need to be isolated. Assuming an ideal self–quarantine, it is possible to forecast the transmission of the disease. Let *t* = 1 be the time the self– quarantine started. In an idealized situation, each person is confined to households. Assuming the average number of persons in a household in Sri Lanka is 5 [3], we applied the classical *SEIR* model to infected households to find the approximate maximum time this household gets infected. According to Figure 10, the number of infected population in a household is less than 1 after 30 days and nearly zero in 60 days. Therefore, we suggest to seal off housed with exposed individuals or villages for minimum 30 days to maximum 60 days duration.

**Fig. 10:**
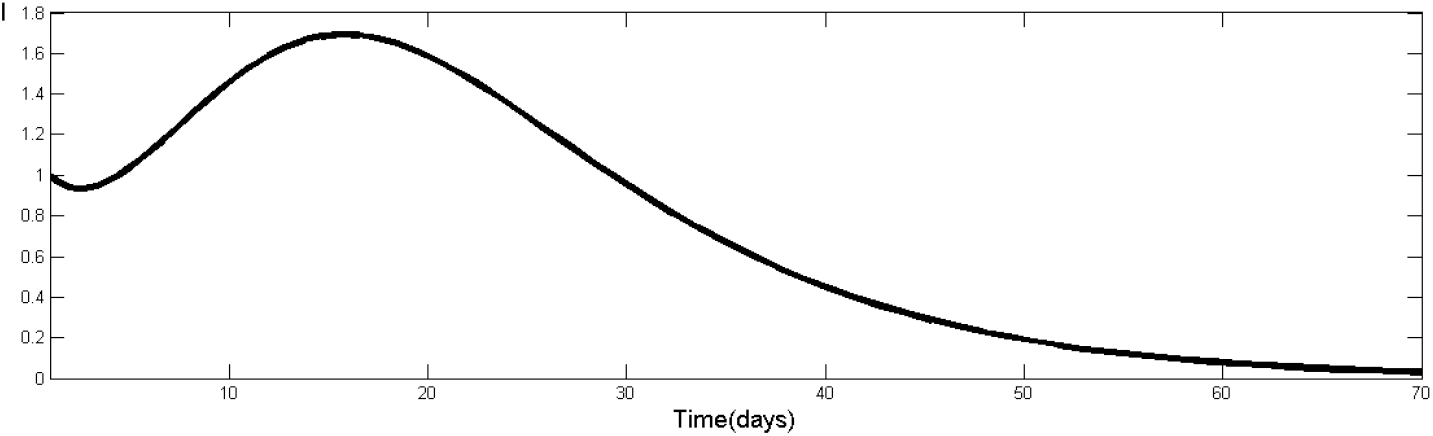
Estimated infected population in household (assuming the average number of persons in a Sri Lankan household to be 5)

In addition, our computational results on the potential consequences of lifting the travel restrictions indicate the possibility the disease being transmitted to some provinces where yet only a few infected persons have been reported. Figure 11 illustrates the impact of curfew relaxation on each province in Sri Lanka as of 19^th^ April and the provinces in Sri Lanka with their closeness centrality measures are presented in Table 2. Comparing the total cases upto 31^th^ May with the rank values, it can be observed that other than the Southern province, all other provinces follow the closeness centrality ranks. A similar analysis with inter–district travel data would provide more insights on how the disease will be transmitted through the transportation network. Thus, the authorities can make decisions on controlling the transport network or allocating provincial or district–wise resources to during a pandemic situation.

**Fig. 11:**
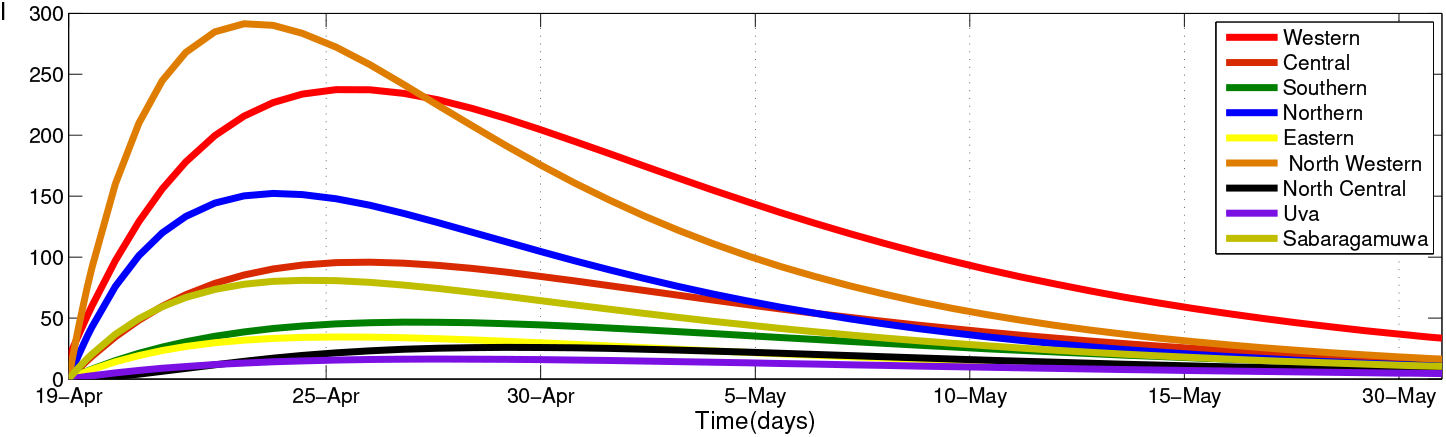
Estimated provincial infected population based on inter–provincial mobility

**Table 2:** Total reported COVID–19 cases (up to 19^th^ April, 2020 and up to 31^st^ May, 2020), normalized closeness centrality results with ranking of the provinces in Sri Lanka

| Province | Total cases<br>(19/04) | Normalized closeness centrality | Rank | Total cases<br>(31/05) |
| --- | --- | --- | --- | --- |
| Western | 154 | 0.1095 | 1 | 220 |
| North Western | 37 | 0.1119 | 2 | 46 |
| Southern | 3 | 0.1861 | 3 | 3 |
| Central | 7 | 0.2314 | 4 | 15 |
| Sabaragamuwa | 8 | 0.2460 | 5 | 9 |
| North Central | 0 | 0.4690 | 6 | 3 |
| Northern | 7 | 0.6267 | 7 | 7 |
| Uva | 1 | 0.7239 | 8 | 5 |
| Eastern | 3 | 1.0000 | 9 | 3 |

## 4 Conclusion

The government of Sri Lanka has implemented a sequence of control measures including nationwide curfew, inter–district travel restriction and lockdown high–risk villages to reduce the growth rate of the COVID–19 pandemic. In this work, we proposed a conceptual model based on classical SEI*R mode*l to describe and analyze the effectiveness of the control strategies implemented in Sri Lanka under five different stages.

The computational results over several contact rate values suggest that reduction of contact rate by not less than 50% would make the outbreak easier to control and the results over several quarantine percentage values suggest that tracing not less than 40% of the contact history of infected individual and quarantined them could contribute to reducing the overall size of COVID–19 outbreak. Assuming an ideal situation for self–quarantine, we implemented the simple *SEIR* model over household data to estimate the approximate time period for the quarantine process and the results indicated the approximate maximum time the household should quarantined is 60 days. In addition, the human mobility inside the country has to be regulated or restricted, in order to stop the disease being transmitted.

## 5 Data Availability

COVID–19 data can be retrieved via Epidemiology Unit, Ministry ofHealth, Sri Lanka, Available at: *http://www.epid.gov.lk/web/index.php?option=com_content&view=article&id=225&Itemid=518&lang=en* and mobility data can be retrieved via National Transport Commission, Sri Lanka, Available at: *https://www.ntc.gov.lk/corporate/pdf/statistics%202015.pdf*

## 6 Conflicts of Interest

The authors declare that they have no conflicts of interest.

## 7 Funding Statement

This work was partly supported by the National Science Foundation grant number RPHS/ 2016/D/05.

## Data Availability

Used publicly available COVID -19 reported data in Sri Lanka

http://www.epid.gov.lk/web/index.php?option=com_content&view=article&id=225&Itemid=518&lang=en

## Notes

### Competing Interest Statement

The authors have declared no competing interest.

